# Psychiatric Symptoms Related to the COVID-19 Pandemic

**DOI:** 10.1101/2020.04.16.20067744

**Authors:** Christopher Rohde, Oskar Hougaard Jefsen, Bettina Nørremark, Andreas Aalkjær Danielsen, Søren Dinesen Østergaard

## Abstract

Individuals who live with mental disorders may be particularly vulnerable to the psychological stress associated with the COVID-19 pandemic. Here, based on screening of 11,072 clinical notes containing a COVID-19 related word (“corona“, “COVID”, “virus”, “epidemic”, “pandemic”, and “contaminate/contamination”), we report data on 918 adult patients from a psychiatric treatment setting in Denmark that display psychiatric symptoms (especially anxiety), which appear to be related to the pandemic and/or its consequential societal changes. This finding underlines the necessity of taking urgent action to mitigate the negative effects of the COVID-19 pandemic on individuals with mental disorders.

## Background

The COVID-19 pandemic and the consequential societal changes are likely to have major health consequences way beyond those caused by the virus infection (1). One of the medical fields that may experience significant consequences of the pandemic is that of psychiatry (2-4). Importantly, individuals who already live with mental disorders may be particularly vulnerable to the psychological stress associated with the pandemic (4, 5). However, data on how the COVID-19 pandemic affects this group of people is largely absent from the literature. Here, we report data from an investigation conducted at the psychiatric services of the Central Denmark Region (CDR), which has a catchment area of approximately 1.3 million people covered by five psychiatric hospitals providing inpatient and outpatient treatment of all types of mental disorders. Based on clinical observation and evidence of the psychological effects of both the current (6) and past (2) pandemics, we expected to find examples of “pandemic-related” symptoms of anxiety, obsessive-compulsive disorder, depression and psychotic disorders (e.g. schizophrenia).

## Methods

We extracted all clinical notes from the period from February 1^st^ to March 23^rd^ 2020 (first verified case of COVID-19 in Denmark was reported on February 26^th^ 2020) (7) from the electronic medical record system used by the psychiatric services of the CDR for individuals (patients) aged 18 or above. In this period there were 412,804 clinical notes reporting on a total of 14,561 adult patients in the psychiatric services of the CDR. Subsequently, we conducted an electronic search for clinical notes containing at least one of the following words: “corona“, “COVID”, “virus”, “epidemic”, “pandemic”, and “contaminate/contamination” (including compound words). A total of 11,072 clinical notes contained at least one of these six pandemic-related words. Then, these 11,072 clinical notes were screened manually (5514 screened by CR and 5558 screened by OHJ) to determine whether they described symptoms that seemed plausibly related to the COVID-19 pandemic. Specifically, the clinical notes were labeled with “0” (no pandemic-related psychiatric symptoms), “1” (possible pandemic-related psychiatric symptom) or “2” (pandemic-related psychiatric symptom). All clinical notes labeled 1 were reassessed by both CR and OHJ and relabeled as either 0 or 2 (lack of consensus was resolved by discussion and/or consultation with SDØ). Throughout this process, it was attempted to distinguish between reactions to the pandemic, which appeared rational and reactions, which seemed pathological. This distinction relied on the use of qualifiers (e.g. “very worried” vs. “somewhat worried”), descriptions of worsening of otherwise stable psychopathology (e.g. “more hopelessness than usual” in a patient with depression), and was based on available knowledge of other patient characteristics (e.g. worries about potential COVID-19 contamination in a patient with severe asthma (not pathological) vs. in a patient with obsessive-compulsive disorder (pathological)). The screening and labeling process outlined above is illustrated in Supplementary Figure 1. After the labeling of all the clinical notes as either 0 (no pandemic-related psychiatric symptoms) or 2 (pandemic-related psychiatric symptom), the latter were coded according to the most dominant psychopathology (see details in the Supplementary Material).

**Figure 1.**
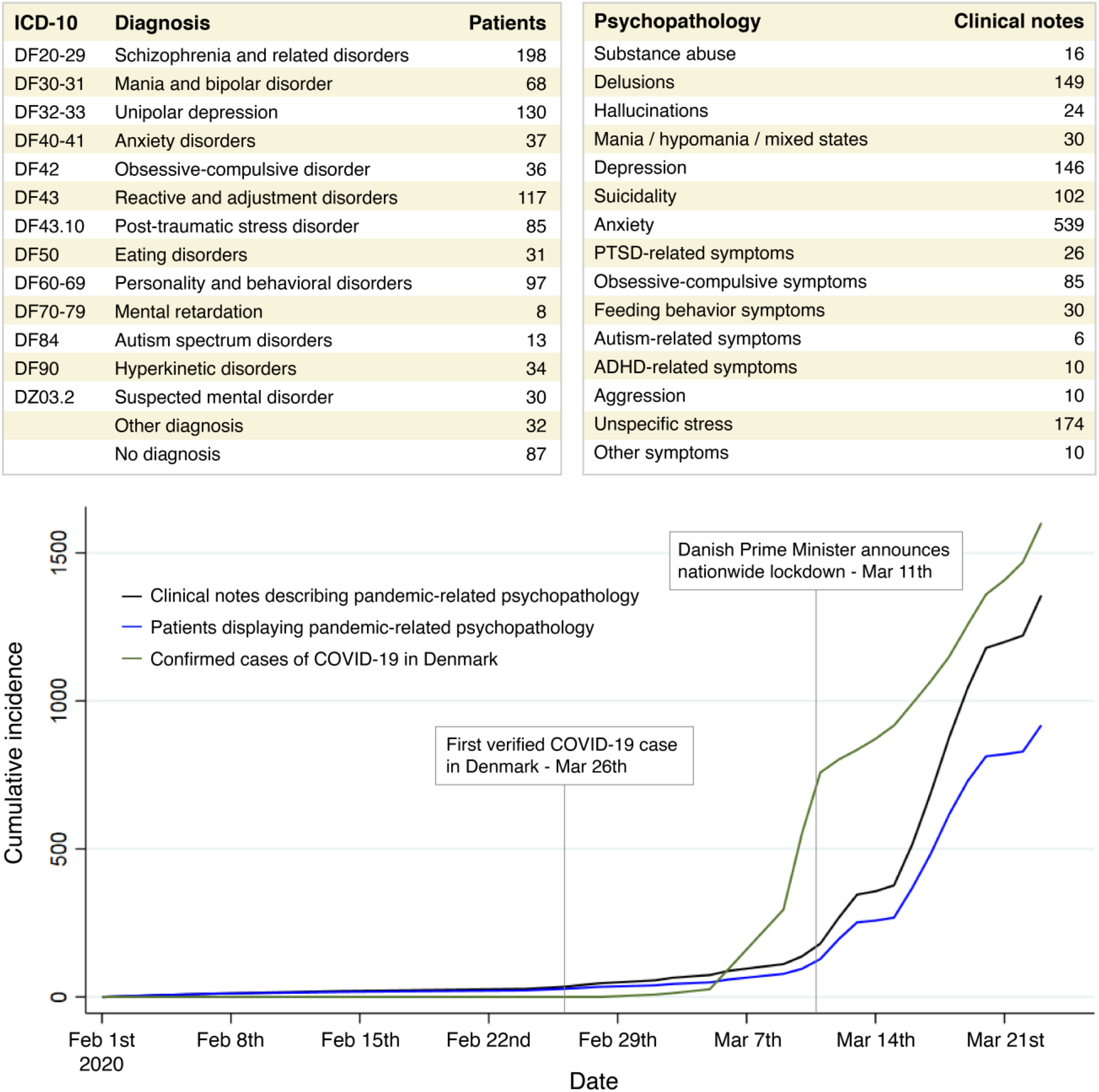
The cumulative incidence of COVID-19 pandemic-related psychopathology among patients with mental disorders in the Central Denmark Region. The nationwide lockdown announced on March 11th 2020 involved closure of kindergartens, schools, teaching institutions, restaurants, bars and many other small businesses. On March 23^rd^ 2020, the lockdown was extended to April 14^th^ 2020. ICD-10: International Classification of Diseases - 10th Revision.

This investigation was approved by the Chief Medical Officer of Psychiatry in the CDR as part of a quality development project (“COVID-19 and mental disorders”), which aims at optimizing the detection and care of patients with COVID-19 pandemic-related psychiatric symptoms in the mental health services of the CDR.

## Results

The initial screening resulted in 9405, 695, and 972 clinical notes labeled with 0 (no pandemic-related psychiatric symptoms), 1 (possible pandemic-related psychiatric symptom), or 2 (pandemic-related psychiatric symptom), respectively. The reassessment of the clinical notes initially labeled as 1 resulted in 310 and 385 notes relabeled with 0 and 2, respectively. Consequently, there were 1357 clinical notes from 918 patients (621 females with a mean age of 36.3 years (SD=14.3) and 297 males with a mean age of 40.9 years (SD=13.8)) describing COVID-19 pandemic-related psychiatric symptoms. As expected, the majority of the 9715 clinical notes not describing COVID-19 pandemic-related psychopathology focused on logistics (e.g. “The appointment was rescheduled to a phone consultation due to the risk of COVID-19”).

The main diagnoses of the 918 patients and the type of the COVID-19 pandemic-related psychopathology described in their clinical notes are reported in Figure 1, which also shows the cumulative incidence of i) patients with pandemic-related psychopathology, ii) clinical notes describing pandemic-related psychopathology, and iii) confirmed cases of COVID-19 in Denmark^5^ in the period from February 1^st^ to March 23^rd^. The figure shows a clear association between the number of COVID-19 cases in Denmark (and the societal restrictions) and the degree of pandemic-related psychopathology (predominantly anxiety).

## Discussion

To our knowledge, this is the first investigation of COVID-19 pandemic-related psychopathology from a large psychiatric treatment setting. The results clearly suggest that this phenomenon exists among individuals with mental illness (2-4). However, as the data from this investigation stems from standard clinical practice where the patients were not systematically assessed for pandemic-related psychopathology, we cannot speak to its overall prevalence. Nevertheless, we believe that our findings underline the necessity of taking urgent action to mitigate the negative effects of the COVID-19 pandemic on individuals with mental disorders. A low-cost intervention would be to simply discuss the ongoing pandemic and its societal consequences with patients seeking care in psychiatric settings - as this may lead to relief of pandemic-related symptoms via general comforting, and clarification of misunderstandings/false beliefs regarding the pandemic. This is also the advice we have provided to our colleagues in the CDR based on these findings. As an example, we have phrased three questions that can be used by the staff to initiate a conversation on the COVID-19 pandemic with their patients:

–*“How do you feel regarding all the things that are currently happening with regard to the coronavirus?”*
–*“Worrying comes easily these days due to the focus on the coronavirus. Is that something your are experiencing too?”*
–*“Many things are different these days due to the coronavirus and it can be difficult to maintain a routine - and do things that one enjoys. How are you doing in that regard?”*

It is our hope that this initiative will aid our colleagues in detecting and addressing any COVID-19 pandemic-related symptoms that their patients may be experiencing.

## Data Availability

Due to the sensitive nature of the data, it is only accessible for quality development projects to employees in the CDR upon application to (and approval by) the Chief Medical Officer of Psychiatry.

## Supplementary material

